# Rapid Access Atrial Fibrillation Clinics in Australia - Modelling outcomes and cost effectiveness

**DOI:** 10.1101/2025.08.08.25333330

**Authors:** Adam C. Livori, Ansu Alex, Tamrat Abebe, J. Simon Bell, Zanfina Ademi, Jedidiah I. Morton

## Abstract

**Aims:** Stroke prevention in patients with atrial fibrillation (AF) requires both estimation of risk and the initiation of anticoagulation treatment where indicated. Rapid access atrial fibrillation (RAAF) clinics are an accepted model of multidisciplinary care to reduce time at risk of stroke, but clinical outcomes and cost-effectiveness of them are uncertain. This study aimed to perform a cost-effectiveness evaluation of a RAAF clinic within a large regional health service in Australia.

**Methods:** We developed a microsimulation model using a cohort of 274 individuals referred to the RAAF clinic between 2022-2023. Clinic data was used to determine risk of stroke, major bleeding, and death from the GARFIELD equation. A comparator was designed by duplicating the cohort and changing the time from referral to consultation to a general cardiology clinic within the same health service (i.e. standard of care). The model ran in daily cycles over a two-year time horizon, with individuals replicated 1,000 times from an initial cohort of 274. The outcomes were strokes, bleeding events, quality-adjusted life years (QALY) and healthcare costs for the RAAF compared to standard of care, which were used to determine incremental cost-effectiveness ratios (ICER), with 5% annual discounting

**Results:** The RAAF clinic participants experienced fewer strokes (5,198 vs 5,303), bleeding events (5,369 vs 5,491) and deaths (14,158 vs 14,413). There were marginal increases in QALYs gained (1.67 vs. 1.66 QALY/person), and cost savings of $74 per person ($14,187 vs $14,261), resulting in a dominant ICER. The ICER remained dominant across one-way and probabilistic sensitivity analyses.

**Conclusion:** RAAF clinics are likely to prevent strokes, bleeding, and are cost-saving and could lead to returns on investment. Adoption of this model of care by policy makers can ensure the delivery of safe, effective and cost-saving care that reduces stroke, bleeding, and death in people with atrial fibrillation.

## Background

Atrial fibrillation (AF) is one of the most common cardiovascular conditions, with an estimated 59.7 million people with AF worldwide [1]. In high income countries, the prevalence of AF is increasing, mainly due to ageing and survival from other cardiovascular diseases such as myocardial infarction [1]. The clinical consequences of AF include the development of heart failure, ischaemic heart disease, and stroke. Cardioembolic stroke caused by AF represents more than 25% of all strokes in Australia [2]. Stroke is associated with increased risks of mortality and morbidity, with 6.5 million deaths and 143 million disability-adjusted life years lost globally in 2019 [3]. The total healthcare expenditure to address the acute and chronic costs of ischaemic stroke is estimated at $47.7 billion dollars in Australia over the next 20 years [3]. Given the prevalence of AF and the health and economic consequences of stroke, interventions to reduce stroke risk in AF are imperative [4].

Clinical guidelines for the management of AF emphasise the importance of minimising the time to therapy optimisation in the prevention of stroke [1, 5, 6]. This means that tools to identify those at risk need to be performed quickly and easily, as well as starting appropriate therapy [7]. This is reflected in consumer-centred research with stroke prevention rated as the most important aspect of AF management, even above prevention of death [8]. The mainstay therapy used in the prevention of stroke is oral anticoagulation (OAC), with the introduction of non-vitamin K oral anticoagulants (NOAC) providing a safer and easier-to-use alternative to warfarin [9]. Meta-analysis of real-world data demonstrated the incidence of OAC prescription in newly diagnosed atrial fibrillation has increased worldwide from 43% in 2010 [95% confidence interval (CI) 22–65%] to 78% in 2018 (95% CI 77–78%) [9].

The challenge is how to ensure that we continue to optimise the use of stroke prevention strategies despite the increase in AF incidence [4]. Several models of care have been shown to enhance both the identification and treatment of people at risk. These models typically involve a combination of multidisciplinary teams (such as cardiologists, pharmacists, and nurses), rapid referral and access streams, and assessment of stroke risk with OAC prescription [10–13]. All of these models represent different approaches to the same problem; however, widespread implementation and ongoing funding for them will require demonstration of cost-effectiveness [14].

The Grampians Health Rapid Access Atrial Fibrillation (RAAF) clinic was established in 2022 at a regional health service to reduce waiting times for stroke risk assessment and treatment of AF [12]. Briefly, the RAAF clinic uses a pharmacist-physician model of care, with people referred to the service first seen by a pharmacist via telehealth to assess stroke risk and, where indicated, begin anticoagulation therapy. The RAAF clinic had a median wait time of 14 days from referral to consultation, considerably less than the median of 224 in general cardiology within the same health service, while meeting all known quality indicators for AF care [12]. No health economic analysis of this intervention has been conducted. Indeed, a systematic review of health economic evaluations to reduce stroke through the use of anticoagulation in AF noted a lack of cost-effectiveness analyses of similar interventions [10]. Moreover, it is unclear how impactful these clinics are on long-term clinical outcomes. Therefore, to address these gaps in the literature, we conducted a cost-effectiveness analysis of the Grampians Health RAAF clinic that aimed to project the impact of the service on stroke, major bleeding, and all-cause mortality over a 2-year period.

## Methods

### Model overview

We designed a microsimulation model using a combination of data sources (Table 1). Inputs were primarily drawn from the evaluation of the RAAF clinic which operated from April 2022 to November 2023 [12], as well as published literature. The model took individuals who attended the clinic and modelled incident stroke, major bleeding, and all-cause mortality in one-day cycles over a 2-year time horizon. The choice of one-day cycles was made to be able to examine small differences in risk and changes in OAC treatment as close to a continuous variable as possible [15]. Given individuals can be without anticoagulation on day 13 (due to being referred to clinic without anticoagulation) and be prescribed it on day 14 (when then attend clinic), this allowed us to adjust stroke, bleeding, death risks accordingly [15]. This is justified by how quickly these medications illicit their anticoagulation effects, thereby changing the risk of stroke and bleeding [1].Full details, including analysis syntax in Stata version 18 (StataCorp, USA), are available in the protocol located a https://github.com/cardiopharmnerd/raafcea. Ethics approval was obtained from the Grampians Health Ballarat Human Research Ethics Committee (Project number107482 LNR//BHSSJOG) and Monash University Human Research Ethics Committee (43967).

**Table 1:** Model Inputs.

| Input | Value | Distribution | Source |
| --- | --- | --- | --- |
| Cohort |  |  |  |
| Demographics | Sex, age, and race | Fixed | RAAF clinic study [12] |
| Risk factors | Weight, pulse, diastolic blood pressure, history of bleeding, heart failure, stroke, kidney disease, coronary artery disease, diabetes, smoking, dementia, concurrent anti-platelets, occlusive carotid disease | Fixed | RAAF clinic study [12] |
| Health states |  |  |  |
| Baseline | Binary of stroke history per individual | Fixed | RAAF clinic study [12] |
| Transition probabilities |  |  |  |
| Stroke | Calculated from GARFIELD equation | Fixed | GARFIELD equation [17] |
| Major bleeding | Calculated from GARFIELD equation | Fixed | GARFIELD equation [17] |
| Death | Calculated from GARFIELD equation | Fixed | GARFIELD equation [17] |
| Acute costs |  |  |  |
| Stroke (first three months) | Age and stroke history specific (mean cost \$36,850) | Gamma | Stroke Foundation data 2024 [21] |
| Major bleeding | \$3,532 (\$2,000;\$7,300) | Gamma | Cohort data from Weber et al [22] |
| Clinic and clinician costs | Specific to general cardiology vs. RAAF (mean cost \$786.31) | Fixed | RAAF clinic study [12] |
| Baseline blood tests | \$110.55 | Fixed | Medicare data [23] |
| Baseline diagnostic tests | \$317.90 | Fixed | Medicare data [23] |
| Chronic costs |  |  |  |
| Stroke (3 months to 2 years) | Age and stroke history specific (mean cost \$15,137) | Gamma | Stroke Foundation data 2024 [21] |
| Medication | Medication specific for each OAC | Fixed | PBS and Ademi et al [24,26] |
| General healthcare | Age and sex specific | Gamma | AIHW [25] |
| Utilities |  |  |  |
| Baseline utility | Individual baseline scores (mean 0.85 (0.81–0.88)) | Fixed | RAAF clinic study [12] |
| Stroke utility | 0.65 (0.61; 0.70) | Beta | Disutility meta-analysis [19] |
| Major bleeding | -0.029 (-0.044; -0.014) | Beta | Disutility analysis from ENGAGE AF-TIMI 48 [18] |

Model entry began at the time of referral to clinic (hospital or community-based doctors), and individuals could enter the model in the health states “alive without previous stroke” or “alive with stroke history” (Figure 1). In each cycle, individuals could remain in their current state, or move from alive without stroke to alive with stroke or have a repeat stroke if they already had stroke history. Individuals could die with or without history of stroke. Major bleeding was a health event rather than a state and could occur multiple times throughout the time horizon. Utilities and costs were tracked for all health states, as well as health events. A schematic of the model is shown in Figure 1, noting multiple strokes and bleeding events were possible. All costs and QALYs were discounted at a rate of 5% per annum [16].

**Figure 1:**
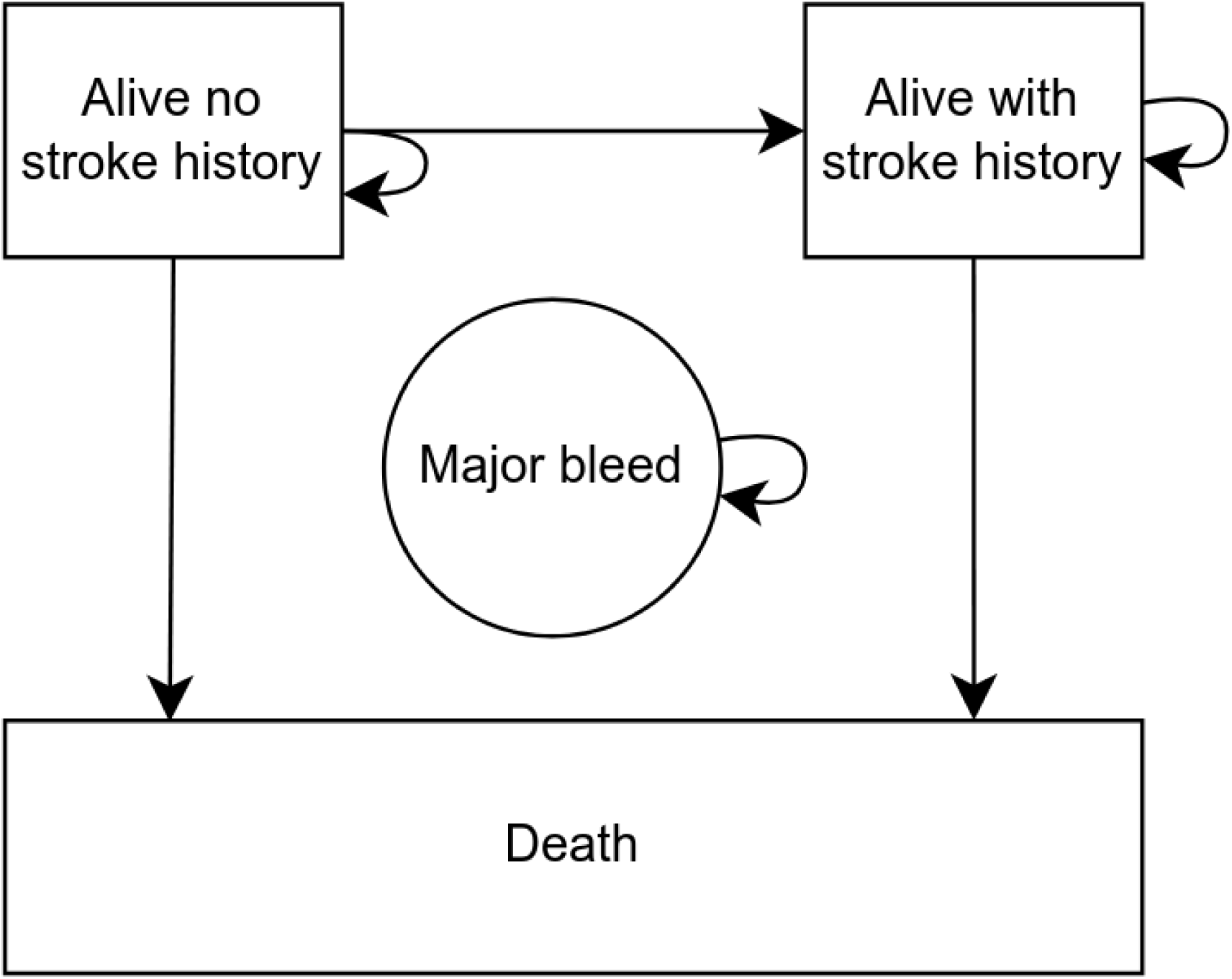
Schematic of the model. Each cycle runs over one day with a two-year time horizon starting from day of referral to the Rapid Access Atrial Fibrillation Clinic. States include alive without stroke, alive with stroke, or dead. Major bleeding events can occur at any cycle while alive.

### Model population

Patient-level data from the RAAF clinic evaluation was used for this cost-effectiveness model [12]. There were 274 individuals, of whom 48% were female (131/274) with a mean age of 68, and 14% (37/274) had a history of stroke and 7% (18/274) had a history of major bleeding (Table 2).Because the RAAF clinic implementation lacked an active comparator, we constructed a comparator that consisting of the same indivudals seen in the RAAF clinic. We kept all predictors of stroke, bleeding and death the same, and only changed their time from referral to consult to reflect the median time to referral from general cardiology services at the same health service from the same period [12]. This is the service patients would normaly be referred to prior to the RAAF clinic commencing. Therefore, the only differences between the groups in the model was the time it takes for that care to be delivered and the cost differences between cardiologist-only care and the RAAF clinic service.

**Table 2:**
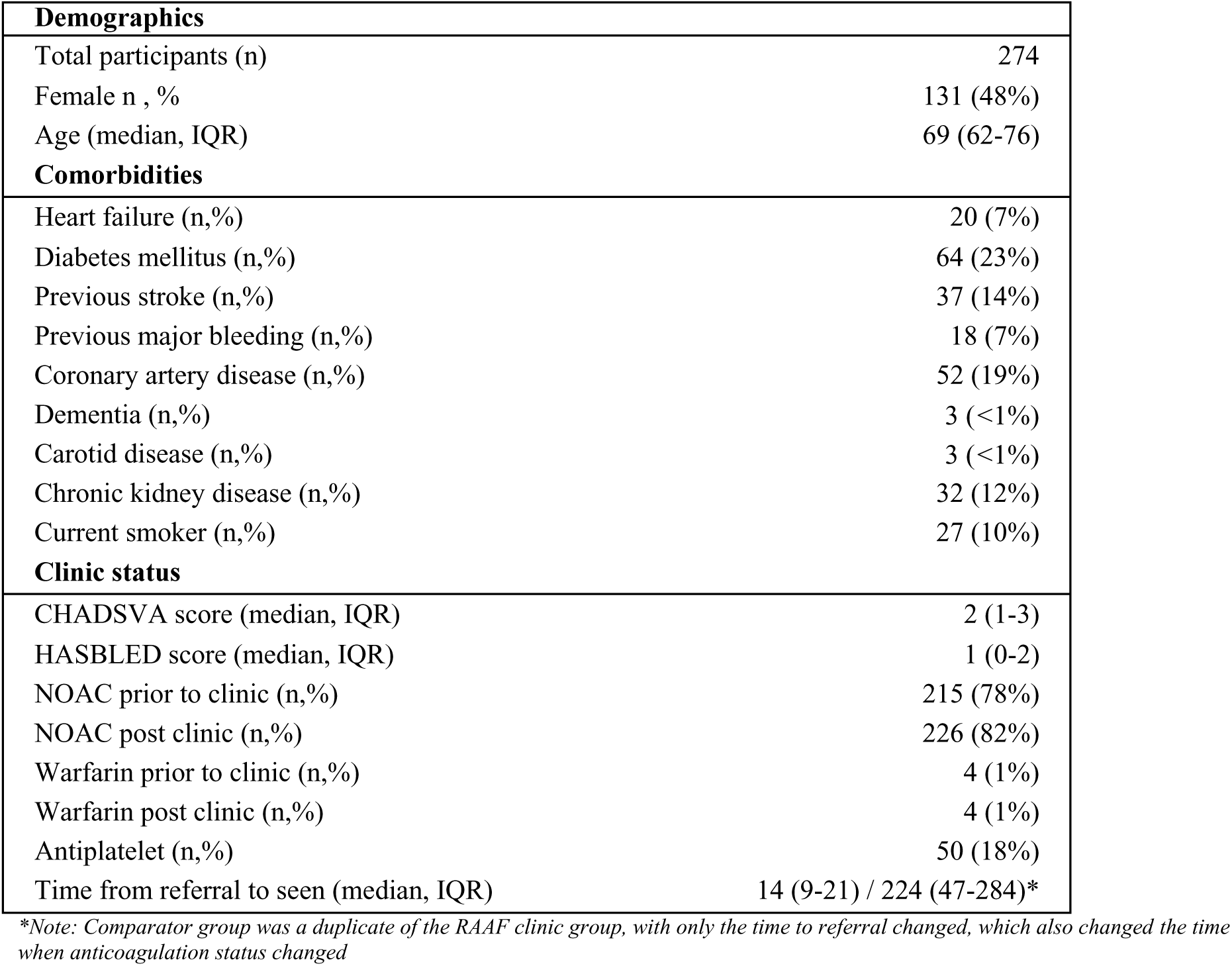
Cohort demographics from the Rapid Access Atrial Fibrillation (RAAF) Clinic.

### Transition probabilities

A 1st order Monte Carlo simulation was completed using patient-level data to predict ischaemic stroke, major bleeding, and all-cause mortality, with risk derived from the GARFIELD equation [17]. The GARFIELD equation was used due to its comprehnsive list of co-variates that include anticoagulation status, and because daily risk calculation is possible. This means that when an individual attends the RAAF clinic, their anticoagulation status may change, and therefore subsequent stroke, bleeding, or death risk also changes [17]. Therefore, the major difference between the likelihood of events occurring with the RAAF and general cardiology clinics is the time spent on anticoagulation.

### Quality of life

Quality of life was assessed using the EuroQol five-dimension questionnaire (EQ-5D) from the RAAF clinic for baseline utility and from published sources for stroke and bleeding disutility [12, 18, 19]. EQ-5D responses were converted into utility scores via a value set that reflect an individual’s perceived health state on a scale where 0 is equivalent to death and 1 is perfect health. The baseline quality of life was taken from the EQ5D-5L questionnaires given to people on referral to the RAAF clinic, and the utility derived from these using an Australian value set [20]. For people with a history of stroke prior to model entry, their baseline utility was assumed to include stroke disutility. For new or recurrent strokes occuring throughout the model, a disutility was applied using a value from a meta-analysis of stroke utility studies [19]. This was done as we did not have follow-up stroke data or repeated responses to EQ5D-5L from individuals seen in the RAAF clinic [19]. This particular utility value was chosen because it provided pooled utility of stroke data specific to embolic stroke averaged over the first two years [19]. This utility value was applied on the day of stroke occurring in the model and continued throughout the time horizon. For example, if the baseline utility was 1.0 and a stroke occurred in cycle 15 with a relative stroke utility of 0.65, then the utility for cycle 15 onwards would be 0.65. Bleeding disutility was derived from a post hoc evaluation of the ENGAGE AF-TIMI trial, and was set at −0.0029 over 270 cycles (9 months) [18]. This study was chosen because it specifically measured the disutility of major bleeding in an AF cohort who used anticoagulation, with a similar multi-morbid cohort. In this instance, if major bleeding occurs in the model, the absolute disutility for major bleeding is subtracted from the individual’s current utility. For example, if the utility at cycle 15 is 0.65 and a bleeding event occurs on cycle 16, then the absolute disutility of −0.029 is applied, resulting in a utility of 0.621 which would continue over 270 cycles (9 months).

Utility values, including disutilities and distributions, are listed in Table 1. The utilities were calculated for each cycle, summed across the time horizon, and divided by 365 to determine the quality-adjusted life years (QALYs) for each individual. The incremental effectiveness was then determined by the difference in QALY between the RAAF clinic and general cardiology.

### Costs

Costs were categorised as acute costs or chronic costs, with all costs in 2024 Australian dollars (AUD;$). Acute stroke costs included the cost of stroke care in the first 3 months and was specific to age and stroke history, derived from the Stroke Foundation of Australia’s Economic Impact of Stroke Report of 2024 [21]. This is the most recent and comprehensive cost data available that specifically focussed on ischaemic stroke costs [21]. These acute costs were applied as a single cost on the day of stroke rather than split over 3 months. This was done because most of these costs are within the hospitalisation period [21].

The acute costs of bleeding events were taken from an Australian study that estimated the cost of different complications of atrial fibrillation in Western Australia [22]. Because major bleeding according to the GARFIELD equation includes both haemorrhagic stroke as well as major gastrointestinal bleeding, we used the average costs for both. As bleeding costs were mainly due to hospital care, it was considered a once-off cost rather than attributed to daily costs over a given period.

Clinic and clinician costs for the RAAF were derived by estimating the clinician time (sourced directly from the RAAF clinic evaluation) and costs from the relative Victorian health professional enterprise bargaining agreements and local health services costs (Supplemental Table 1) [12]. These costs were only applied if the individual was alive on the day of consultation within the model. The costs of baseline blood tests, diagnostic tests, and medication costs were all sourced from the Australian government (Medicare Benefits Schedule (MBS) and Pharmaceutical Benefits Scheme (PBS)) [23, 24]. Similarly to clinic costs, these were only applied if the individual had survived from their referral to consultation time. Medication costs were applied daily provided that the individual was alive, with 100% adherence to therapy assumed for costs (i.e., all medications were dispensed; assumptions of medication efficacy did not rely on 100% adherence). Total anticoagulation costs were the sum of total cost of medication therapy over the time horizon.

Chronic costs of stroke were taken from the same source for acute stroke costs; in the source report, costs were presented as costs 3 months after stroke to 2 years [21]. This value was converted into a daily cost (and assumed equal across the 3 month to 2 year period) for each cycle. If an individual did not have a stroke during the time horizon, general healthcare costs (specific to age and sex) were applied using Australian health cost data from the Australian Institute of Health and Welfare (AIHW) [25].The combination of total QALYs and costs for the RAAF and general cardiology groups was then used to calculate an incremental cost-effectiveness ratio (ICER), defined as intervention costs minus comparator costs, divided by the intervention QALYs minus the comparator QALYS. For the base case, all the point estimates for the inputs listed in Table 1 were used in the model and subsequent ICER calculation. A willingness-to-pay threshold of $50,000 was assumed when reviewing the ICER [26].

### Scenario analyses

The first scenario analysis removed discounting from the model to understand its impact on the ICER. In the second, we compared the RAAF to a hypothetical traditional medical model that reduced time from referral to consultation to 14 days (i.e., the only difference between the RAAF and medical model being which clinician – pharmacist or cardiologist – performs an initial patient evaluation).

### One-way sensitivity analysis

To quantify the effects of uncertainty within each of the inputs listed in Table 1, we repeated the microsimulation and isolated each of the non-fixed inputs: disutility of major bleeding; disutility of stroke; acute cost of stroke; acute cost of major bleeding; chronic cost of stroke; chronic cost of healthcare. We quantified the total QALYs and costs at the lower and upper bound of each input, producing an ICER for each that we presented in a tornado plot.

### Probabilistic sensitivity analysis

In addition to the first-order Monte Carlo simulation used to quantify state transitions and events, we undertook a second-order Monte Carlo microsimulation for a probabilistic sensitivity analysis (PSA). We randomly sampled from the distributions of each non-fixed input in table 1. The end product of the PSA was 1000 individual QALY and costs totals.

## Results

### Base case

Among 274,000 simulated individuals, patients seen in the RAAF clinic, compared with general cardiology, experienced fewer strokes (5,198 vs. 5,303), fewer bleeding events (5,369 vs. 5,494), and fewer deaths (14,158 vs. 14,413) throughout the time horizon (Table 3). This resulted in a marginally larger number of QALYs with a difference of less than 1%. However, due to the lower operating costs for the RAAF clinic and lower rates of stroke and bleeding, there was a cost saving of $74 per person seen in the RAAF clinic, resulting in a dominant ICER (i.e. QALY gain with negative incremental costs).

**Table 3:**
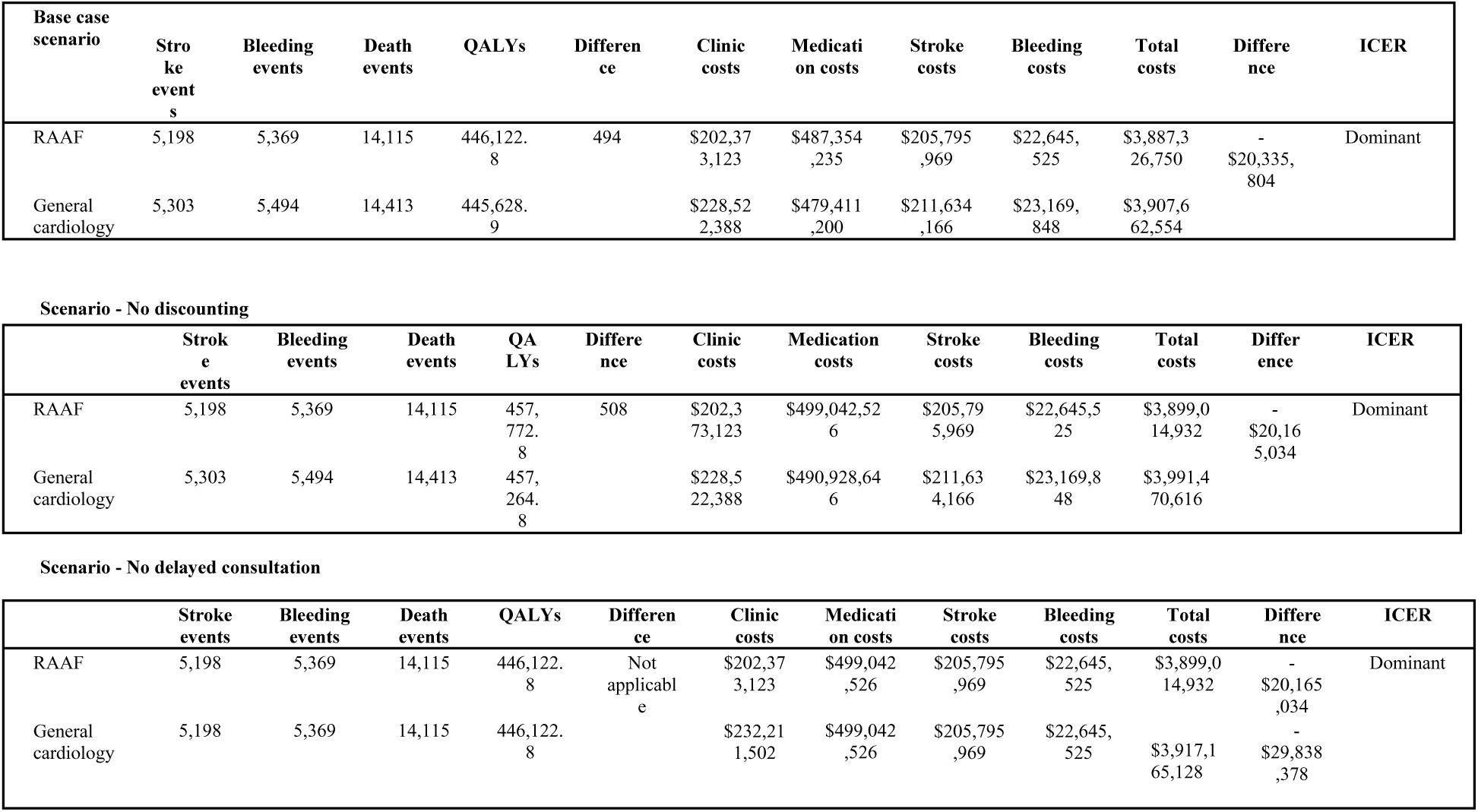
Results from base case and scenario analysis.

### Scenario analyses

There was minimal impact on the ICER when considering the exclusion of discounting for the first scenario. The second scenario considered how the general cardiology service would compare if it could achieve the same time from referral to consultation and demonstrated a $104 saving per person.

### Sensitivity analyses

The tornado plot in Figure 2 demonstrates the impact of changing individual input values to their upper and lower limits. The most influential parameter was chronic cost of healthcare, with a change in the ICER of $857 below and $857 above; although no input had any material impact on the ICER, which remained dominant in all analyses. The smallest difference was present in the disutility for bleeding, with impacts of $143 below and $144 above the base case ICER. Overall, all possible input values in the one-way deterministic sensitivity analyses still demonstrate that the RAAF clinic is a dominant strategy.

**Figure 2:**
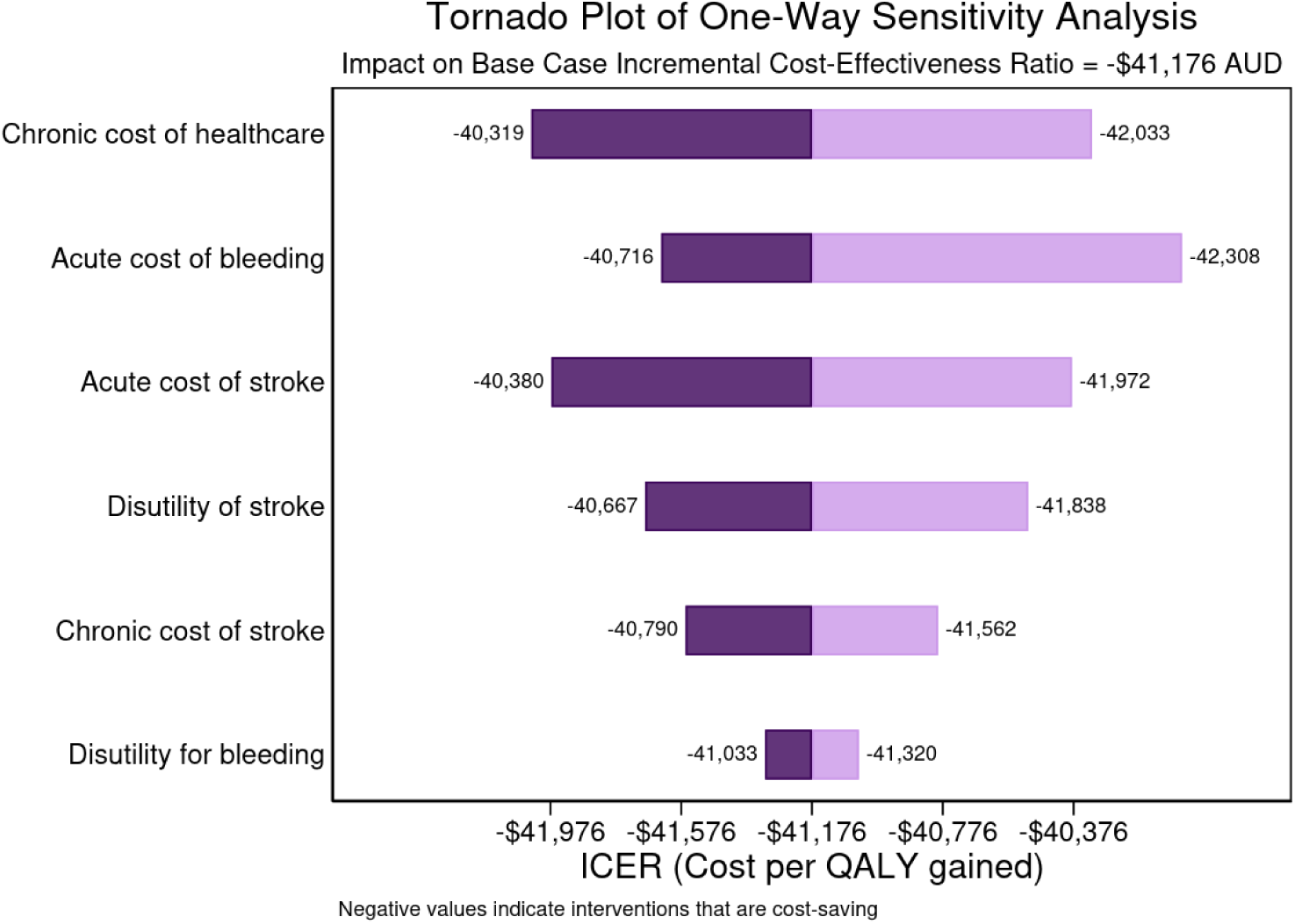
Tornado diagram for one-way sensitivity analysis. This demonstrates the uncertainty in the calculated incremental cost-effectiveness ratio (ICER) for the model’s inputs. Lower limit ICERs reflect the model when the lower limit of the estimate is used, and upper limit ICERs when the upper limit is used. Illustrated ICERs are relative to the base case value of −$41,176 per quality-adjusted life year

From a healthcare perspective, all 1000 model simulations demonstrated that the RAAF clinic was a dominant strategy compared to general cardiology (Figure 3). This means that in all possible iterations of the model, it produced dominant alternative for the management of AF.

**Figure 3:**
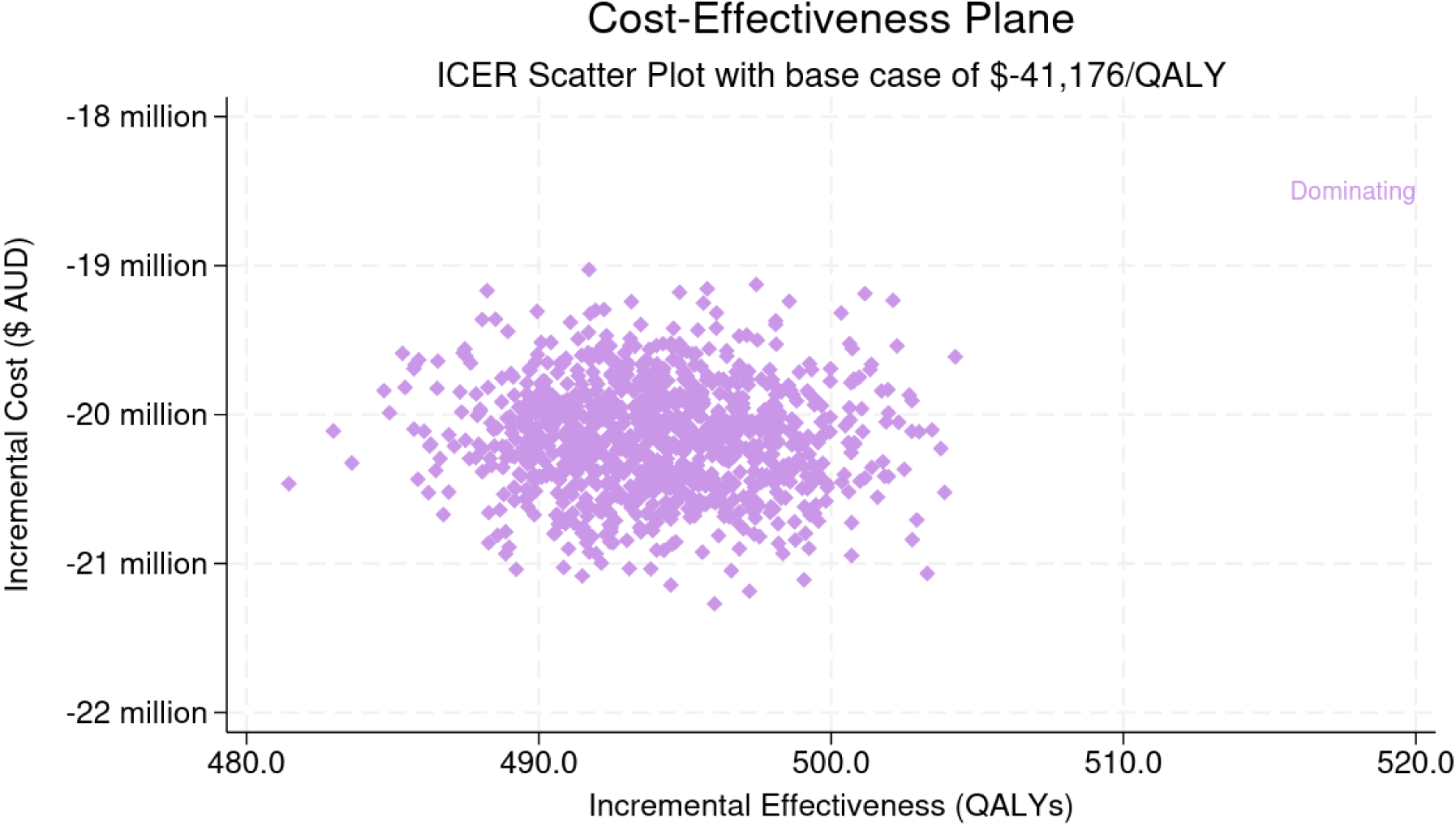
Scatter plot of results from probabilistic sensitivity analysis. The results of the 1000 simulations are plotted, with all resulting ICERs within the dominant region of the cost-effectiveness plane which includes the base case value of −$41,176 per quality-adjusted life year

## Discussion

### Main findings

In this study, we showed that the implementation of a RAAF clinic within a large regional health service in Australia would be an effective and cost-saving (dominant) strategy from a healthcare perspective. The RAAF clinic demonstrated gains in QALYs attributable to a slightly reduced incidence of stroke (5,198 vs. 5303), reduced bleeding events (5,369 vs. 5,494), and all-cause death (14,115 vs. 14,413). These results support the implementation of multidisciplinary clinics that reduce the time at risk for stroke among people with atrial fibrillation.

### Time to review and outcome reduction

In our model, death, stroke and bleeding risk was determined by the GARFIELD equation, and the only variable within the equation we modified was whether anticoagulation was present or not [17]. For death and stroke, the presence of OAC reduces stroke risk, and so given we increased anticoagulation in those at risk of stroke earlier in the RAAF clinic, as there were less strokes and deaths. When we consider bleeding, the presence of OAC increases risk, yet we also saw a reduction in bleeding events with the RAAF clinic. This finding was also noted in the RAAF clinic evaluation, because there were many patients on OAC at the time of referral that did not need to be [12]. Indeed, those with minimal stroke risk (CHADSVA of 0) on OAC at the time of referral were 41% (13/32) prior to RAAF clinic and was reduced to 19% (6/32) following consultation [12]. The competing risk of bleeding is something that is noted in many clinical trials of OAC, but can often be missing from anticoagulation based service evaluations [1, 11, 13]. Therefore, given that stroke, death, and bleeding are all time-dependent risks that are altered by the presence of OAC, faster assessment and optimisation of OAC use will reduce risk and thus reduce events. Comparison with other models of care

Cost-effectiveness studies of pharmacist interventions have been previously summarised in a systematic review by Elliot et al [27]. They noted the inherent complexity of being able to draw conclusions due to methodological differences and a lack of measures, such as utilities, so that ICERs can be calculated. Our study built on these limitations by considering utility, using as many inputs derived from the intervention as possible, and limiting assumptions on the comparator [27]. We intentionally wanted to ‘fix’ the effects of the RAAF clinic solely on time to assessment, rather than making further assumptions on whether the model offered more or less benefit to other aspects of patient care. These multidisciplinary models of care are encouraged by international guidelines and supported by high-level evidence [1, 28]. Our positive results for cost effectiveness are similar to that of a cost-effectiveness analysis of the REACH clinical trial (ICER −$7,636 per QALY), where pharmacist consults were introduced in addition to standard care to reduce cardiovascular events [29]. The authors commented that the reduced cost of pharmacist care combined with an earlier intervention on time-dependent risk were the drivers of the dominant strategy, which is consistent with our work [29].

A mixed study systematic review of nurse-led atrial fibrillation clinics drew similar conclusions to that of Elliot et al, reporting shorter wait times, cost-effective interventions, and comparable quality of care to standard medical care [30]. The authors noted that isolating individual professional contributions is difficult and that none of the studies considered regional and rural applications of care interventions [30]. Our work builds on this research by focussing on a regional health setting and considering a collaborative model of care rather than isolating individual contributions from health professionals [30].

### Limitations

There were several limitations to our study. Firstly, the lack of an active comparator as part of the original RAAF clinic evaluation means that we are not necessarily seeing the actual differences in time from referral to consultation for a separate but similarly matched cohort. This limits us to understanding the true differences between care due to having a simulated comparator [10]. Secondly, we assumed that the stroke risk over the year is the sum of 365 individual days of risk. This assumes that each day’s risk is independent and identical. We know that each day of a person’s life is not independent, and we are giving an average daily risk under the assumption of a uniform distribution [31]. Third, we took a conservative approach in assuming 100% adherence to OAC therapy for costs of medications, however the GARFIELD equation used self-reported adherence for its risk calculation from real-world data, which may not reflect 100% adherence [10]. A meta-analysis of studies of real-world adherence of 594,784 individuals demonstrated a mean proportion of days covered of 77% (95% CI, 75–80%) [32]. However, we know that adherence definitions are difficult to define, and transparency in reporting proportion of days covered is lacking [33, 34]. Individual-level adherence measures within the RAAF clinic may have provided a better representation of daily risks. Finally, our study was limited to one health service within a regional area of Victoria, Australia. Differences in AF care availability and in particular, access and waiting times for specialist care, may reduce the generalisability of our findings.

### Implications

The RAAF clinic is a cost-saving intervention even when the reductions in wait times are modelled with standard care. These findings support the creation of clinics throughout the health system to address stroke risk in people with AF where the time from referral to consultation is prolonged. Adoption of the model should include measurement of baseline and subsequent patient reported quality of life measures, as well as methods for addressing adherence to therapies and active comparison with other models of care [10, 32]. Given that stroke and major bleeding are both predictable and preventable conditions, future studies should also consider the impacts on productivity of the intervention [35]. The use of more novel measures, such as productivity-adjusted life years, will facilitate a broader comparison of the impact of the RAAF clinic for health services, policy makers, and society [35].

## Conclusion

The RAAF clinic was shown to be effective and cost-saving, a finding which was consistent across multiple sensitivity and scenario analyses. This study offers a high degree of support for multi-disciplinary clinics designed to reduce the time at risk for stroke for people with atrial fibrillation. We recommend that service providers and policy makers implement these models of care to reduce stroke risk among a growing population of people with AF and the concomitant increasing burden on healthcare spending.

## Declaration of interest

- AL Consulting for Sanofi, Boehringer Ingelheim, Novartis
- JSB is supported by a National Health and Medical Research Council (NHMRC) Boosting Dementia Research Leadership Fellowship and has received grant funding or consulting funds from the NHMRC, Medical Research Future Fund, Victorian Government Department of Health, Dementia Australia Research Foundation, Yulgilbar Foundation, Aged Care Quality and Safety Commission, Dementia Centre for Research Collaboration, Pharmaceutical Society of Australia, Society of Hospital Pharmacists of Australia, GlaxoSmithKline Supported Studies Programme, Amgen, and several aged care provider organizations unrelated to this work. All grants and consulting funds were paid to the employing institution.
- AA, TA, ZA, JIM No conflicts to declare relevant to this publication

## Acknowledgments

The authors acknowledge the project management support provided by Safer Care Victoria and the Cardiovascular Learning Health Network. The authors also acknowledge the Internal Medical Services and Pharmacy departments of Grampians Health for approving the implementation and subsequent ongoing funding for this model of care.

## Data Availability

The individual-level data underlying this article cannot be shared publicly due to privacy reasons. However, the protocol containing all code used to generate this study is available at: https://github.com/cardiopharmnerd/raafcea.

## Funding

JIM was supported by a Heart Foundation Postdoctoral Fellowship (ID: 108269-2024) from the National Heart Foundation of Australia.

## CRediT authorship statement

**Adam C. Livori**: Writing – original draft, Project administration, Methodology, Investigation, Formal analysis, Data curation, Conceptualization. **Ansu Alex**: Writing – review & editing, Data curation. **Tamrat Abebe**: Writing – review & editing, Data curation. **J. Simon Bell**: Writing – review & editing, Supervision, Resources. **Zanfina Ademi:** Writing – review & editing, Supervision, Formal analysis. **Jedidiah I. Morton**: Writing – review & editing, Visualization, Validation, vision, Methodology, Formal analysis, Conceptualization.

## Conflicts of Interest

AL Consulting for Sanofi, Boehringer Ingelheim, Novartis.

AA, TA, ZA, JIM No conflicts to declare relevant to this publication.

JSB is supported by a National Health and Medical Research Council (NHMRC) Boosting Dementia Research Leadership Fellowship and has received grant funding or consulting funds from the NHMRC, Medical Research Future Fund, Victorian Government Department of Health, Dementia Australia Research Foundation, Yulgilbar Foundation, Aged Care Quality and Safety Commission, Dementia Centre for Research Collaboration, Pharmaceutical Society of Australia, Society of Hospital Pharmacists of Australia, GlaxoSmithKline Supported Studies Programme, Amgen, and several aged care provider organizations unrelated to this work. All grants and consulting funds were paid to the employing institution.

## Notes

### Clinical Trial

N/A

### Author Declarations

Ethics approval was obtained from the Grampians Health Ballarat Human Research Ethics Committee (Project number107482 LNR//BHSSJOG) and Monash University Human Research Ethics Committee (43967).

## References

1. Van Gelder IC, Rienstra M, Bunting KV, et al. 2024 ESC Guidelines for the management of atrial fibrillation developed in collaboration with the European Association for Cardio-Thoracic Surgery (EACTS). European Heart Journal 2024.

2. Australian Institute of Health and Welfare. Heart, stroke and vascular disease: Australian facts. Tech. rep. Canberra: Australian Institute of Health and Welfare, 2023.

3. Abebe TB, Ilomaki J, Livori A, Bell JS, Morton JI, and Ademi Z. Current and Future Cost Burden of Ischemic Stroke in Australia: Dynamic Model. Neuroepidemiology 2024.

4. Lippi G, Sanchis-Gomar F, and Cervellin G. Global epidemiology of atrial fibrillation: An increasing epidemic and public health challenge. International Journal of Stroke 2021;16:217– 21.

5. Joglar JA, Chung MK, Armbruster AL, et al. 2023 ACC/AHA/ACCP/HRS Guideline for the Diagnosis and Management of Atrial Fibrillation. Journal of the American College of Cardiology 2023.

6. Brieger D, Amerena J, Attia J, et al. National Heart Foundation of Australia and the Cardiac Society of Australia and New Zealand: Australian Clinical Guidelines for the Diagnosis and Management of Atrial Fibrillation 2018. Heart Lung Circ 2018;27:1209–66.

7. Arbelo E, Aktaa S, Bollmann A, et al. Quality indicators for the care and outcomes of adults with atrial fibrillation. Europace 2021;23:494–5.

8. Swart EC, Good CB, Henderson R, et al. Identifying outcome measures for atrial fibrillation value-based contracting using the Delphi method. Research in Social and Administrative Pharmacy 2022;18:3425–31.

9. Grymonprez M, Simoens C, Steurbaut S, De Backer TL, and Lahousse L. Worldwide trends in oral anticoagulant use in patients with atrial fibrillation from 2010 to 2018: a systematic review and meta-analysis. 2022. DOI: 10.1093/europace/euab303.

10. Jones NR, Crawford W, Yang Y, Hobbs FR, Taylor CJ, and Petrou S. A Systematic Review of Economic Aspects of Service Interventions to Increase Anticoagulation Use in Atrial Fibrillation. Thrombosis and Haemostasis 2022;122:394–405.

11. Klimis H, Von Huben A, Turnbull S, et al. Rapid Access Arrhythmia Clinics (RAACs) Versus Usual Care: Improving Efficiency and Safety of Arrhythmia Management. Heart Lung and Circulation 2021;30:665–73.

12. Livori AC, Kuruppumullage R, Simmons M, et al. Evaluating the implementation of a rapid access atrial fibrillation clinic utilising a pharmacist-physician model of care. Research in Social and Administrative Pharmacy 2025.

13. Gillis AM, Burland L, Arnburg Bscn B, et al. Treating the right patient at the right time: An innovative approach to the management of atrial fibrillation. Tech. rep. 2008.

14. Marquina C, Ademi Z, Zomer E, Ofori-Asenso R, Tate R, and Liew D. Cost Burden and Cost-Effective Analysis of the Nationwide Implementation of the Quality in Acute Stroke Care Protocol in Australia. Journal of Stroke and Cerebrovascular Diseases 2021;30.

15. O’Mahony JF, Newall AT, and Rosmalen J van. Dealing with Time in Health Economic Evaluation: Methodological Issues and Recommendations for Practice. PharmacoEconomics 2015;33:1255–68.

16. Manipis K, Viney R, De Abreu Louren, co R, et al. Health Technology Assessment methods: Economic Evaluation. Australian Health Technology Assessment Methods and Policy Review. Tech. rep. Canberra, Australia: Australian Government, Department of Health and Aged Care., 2023. URL: http://www.chere.uts.edu.au.

17. Fox KA, Virdone S, Pieper KS, et al. GARFIELD-AF risk score for mortality, stroke, and bleeding within 2 years in patients with atrial fibrillation. European Heart Journal - Quality of Care and Clinical Outcomes 2022;8:214–27.

18. Wang K, Li H, Kwong WJ, et al. Impact of spontaneous extracranial bleeding events on health state utility in patients with atrial fibrillation: Results from the ENGAGE AF-TIMI 48 trial. Journal of the American Heart Association 2017;6.

19. Joundi R, Adekanye J, Leung A, et al. Health State Utility Values in People With Stroke: A Systematic Review and Meta-Analysis. Journal of the American Heart Association 2022;11.

20. Norman R, Mulhern B, Lancsar E, et al. The Use of a Discrete Choice Experiment Including Both Duration and Dead for the Development of an EQ-5D-5L Value Set for Australia. PharmacoEconomics 2023;41:427–38.

21. Kim J, Neville E, Dalli L, et al. Economic Impact of Stroke 2024. Tech. rep. Melbourne: Stroke Foundation of Australia, 2024.

22. Weber C, Hung J, Atkins ER, Hickling S, Briffa T, and Li I. Long-Term Pattern and Associated Costs of Re-hospitalisations in Patients After Index Atrial Fibrillation Admission in Western Australia, 2011–2017. Heart Lung and Circulation 2024;33:55–64.

23. Department of Health and Aged Care. Medicare Benefits Schedule (MBS) [Internet]. 2024.

24. Department of Health and Aged Care. Pharmaceutical Benefits Scheme (PBS) [Internet]. 2022.

25. Australian Institute of Health and Welfare. Health system spending on disease and injury. 2023.

26. Wang S, Gum D, and Merlin T. Comparing the ICERs in Medicine Reimbursement Submissions to NICE and PBAC—Does the Presence of an Explicit Threshold Affect the ICER Proposed? Value in Health 2018;21:938–43.

27. Elliott RA, Putman K, Davies J, and Annemans L. A Review of the Methodological Challenges in Assessing the Cost Effectiveness of Pharmacist Interventions. 2014. DOI: 10.1007/s40273-014-0197-z.

28. Livori AC, Prosser A, and Levkovich B. Clinical outcome measures in the assessment of impact of pharmacists in cardiology ambulatory care: A systematic review. Research in Social and Administrative Pharmacy 2023;19:432–44.

29. Al Hamarneh YN, Johnston K, Marra CA, and Tsuyuki RT. Pharmacist prescribing and care improves cardiovascular risk, but is it cost-effective? A cost-effectiveness analysis of the Rx-EACH study. Canadian Pharmacists Journal 2019;152:257–66.

30. Rush KL, Burton L, Schaab K, and Lukey A. The impact of nurse-led atrial fibrillation clinics on patient and healthcare outcomes: a systematic mixed studies review. 2019. DOI: 10.1177/1474515119845198.

31. Harrell FE. Regression Modeling Strategies With Applications to Linear Models, Logistic and Ordinal Regression, and Survival Analysis Second Edition Springer Series in Statistics. Tech. rep. 2015. DOI: 10.1007/978-3-319-19425-7. URL: http://www.springer.com/series/692.

32. Ozaki AF, Choi AS, Le QT, et al. Real-World Adherence and Persistence to Direct Oral Anticoagulants in Patients With Atrial Fibrillation: A Systematic Review and Meta-Analysis. Circulation: Cardiovascular Quality and Outcomes 2020;13:E005969.

33. Livori AC, Dalli L, Nicholls SJ, and Nelson AJ. Defining, measuring, and addressing medication non-adherence in cardiovascular disease. 2024. DOI: 10.1080/14796678.2024.2433888.

34. Dalli LL, Kilkenny MF, Arnet I, et al. Towards better reporting of the proportion of days covered method in cardiovascular medication adherence: A scoping review and new tool TEN-SPIDERS. 2022. DOI: 10.1111/bcp.15391.

35. Ademi Z, Ackerman IN, Zomer E, and Liew D. Productivity-Adjusted Life-Years: A New Metric for Quantifying Disease Burden. 2021. DOI: 10.1007/s40273-020-00999-z.

36. Ademi Z, Pasupathi K, and Liew D. Cost-effectiveness of apixaban compared to warfarin in the management of atrial fibrillation in Australia. European Journal of Preventive Cardiology 2015;22:344–53.

